# Remote Intervention to Support Caregivers of Latino People with Dementia: Feasibility Study

**DOI:** 10.1101/2025.09.18.25336084

**Authors:** Jaime Perales-Puchalt, Mariana Ramírez-Mantilla, Mónica Fracachán-Cabrera, Christina Baker, Jeffrey M. Burns

## Abstract

**Background and objectives:** Latino people with dementia and their caregivers have poor mental health. We tested the feasibility, acceptability, and preliminary efficacy of a texting and phone call intervention to support informal caregivers of Latino people with dementia.

**Research design and methods:** We enrolled 100 caregivers of 84 Latino people with dementia into a one-arm pre-post intervention trial. This caregiver intervention was remote, bilingual, lasted six months and included optional monthly phone calls to address unmet needs, and a bidirectional texting program focused on dementia education, skill-building and community resources. Outcomes included feasibility, acceptability, and preliminary efficacy (e.g., depressive symptomatology) within six months, measured via surveys and metrics of intervention usage.

**Results:** Eighty-eight percent of caregivers completed the follow-up assessment. Four caregivers unsubscribed from the intervention, 69.0% held at least one phone call visit, 91.0% sent at least one text message, 95.5% reported complying with at least some intervention recommendations, and 95.0% reported very or extremely high satisfaction levels. Effect sizes of change in all preliminary efficacy outcomes increased gradually as we restricted samples to those with worse baseline outcome values.

**Discussion:** The intervention demonstrated high levels of acceptability and feasibility. Phone calls did not engage caregivers as much as texting.

**Implications:** Future efficacy studies should restrict their eligibility criteria based on their outcome of interest. Outcomes with most efficacy promise include caregiver distress, depression and life satisfaction, and person with dementia’s depression.

## Introduction

Dementia has a devastating mental health toll on people with dementia (PWD) and their informal caregivers. PWD experience higher levels of most neuropsychiatric symptoms compared to those without dementia (Savva et al., 2009). Informal caregivers of PWD are also more likely than non-caregivers and caregivers of people with other conditions to experience depression and anxiety disorders. For example, the prevalence of depressive disorders among informal caregivers of PWD is 30-40%, compared to 5-17% for non-caregivers of similar ages or caregivers of people with stroke (20%) or schizophrenia (19%) (Atteih et al., 2015; Cuijpers, 2005; Sallim et al., 2015; Thunyadee et al., 2015). The prevalence of anxiety disorders is also higher among informal caregivers of PWD (44%) than caregivers of people with stroke (31%) (Atteih et al., 2015; Sallim et al., 2015).

The mental health toll of dementia disproportionately impacts the Latino community. Compared to non-Latino Whites, Latinos experience a higher dementia risk (Hudomiet et al., 2022), and Latino PWD experience a higher prevalence of most neuropsychiatric symptoms (Hinton et al., 2006; Lyketsos et al., 2000; Salazar et al., 2017; Sink et al., 2004). Latinos are also more likely to become informal caregivers (Friedman et al., 2015) and experience more depression than their non-Latino White peers (Chen et al., 2020).

Numerous dementia care support interventions exist and have shown to improve mental health outcomes among PWD and their caregivers (Walter & Pinquart, 2020). However, Latinos are less likely than non-Latino Whites to access dementia care support (Scharlach et al., 2008). This lower access is potentially a consequence of most studies not centering their interventions on the Latino community (Gilmore-Bykovskyi et al., 2018), as these experience unique that might reduce traditional caregiver support interventions’ feasibility and efficacy, including time constraints, transportation, language, and cultural barriers (Monahan et al., 1992; Scharlach et al., 2008). While most of the 23 existing caregiver support trials reported among Latino caregivers of people with dementia address factors such as language and stigma, other barriers are rarely addressed, which may be impacting their implementation in the real world (Dessy et al., 2022). Limitations of these interventions include a rigid curriculum (Gallagher-Thompson et al., 2015), low accessibility of face-to-face or web-based interventions (Grossman et al., 2018; Kajiyama et al., 2018; Waller et al., 2017), and a substantial reliance on licensed staff (Butler et al., 2020). Therefore, it is urgent to develop efficacious caregiver support interventions that are targeted to Latinos and are easy to access.

To address these barriers Latinos experience, we developed CuidaTEXT, which is, to our knowledge, the first Short Message Service (SMS) texting intervention for caregiver support of PWD (Perales-Puchalt et al., 2021). CuidaTEXT is highly accessible because, despite addressing stigma and cultural factors, nearly all Latinos have access to SMS texting via their phones and can access them irrespective of their location and asynchronously (Duggan, 2013; Pew Research Center, 2021). Our previous feasibility study (n=24) has shown high levels of feasibility, acceptability and preliminary efficacy (Perales-Puchalt, Ramírez-Mantilla, et al., 2022). In the current study, we aim to test the feasibility, acceptability and preliminary efficacy of CuidaTEXT plus optional monthly phone calls among 100 mostly Latino informal caregivers and the Latino PWD they care for. The combination of texting and phone calls may have a stronger impact than purely texting, but phone calls may add access complications, since these require synchronous communication. Compared to our previous feasibility studies, in the current study we expand the focus on PWD outcomes, therefore broadening our knowledge of the potential impact of remote caregiver support interventions. This development corresponds to Stage 1b of the NIH Stage Model for Behavioral Intervention Development (feasibility testing) (Onken et al., 2014).

## Methods

This was a one-arm pre-post-intervention trial that administered surveys at baseline and six months (end of treatment). We have described the methods elsewhere (Perales-Puchalt et al.). We enrolled caregiver-PWD dyad participants from multiple sources from May 2022 to February 2024. Eligibility for both the caregiver and PWD included speaking Spanish or English, ability to understand the informed consent and being 18 or older. Caregivers had to be a relative or friend of the PWD, provide emotional or physical support, be in personal contact with them at least weekly, and own a cellphone with a flat fee for SMS texting. PWD had to identify as Latino and have a clinical or research dementia/mild cognitive impairment diagnosis. The study allowed more than one dyad per person with dementia. All study procedures were approved by the Institutional Review Board of the University of Kansas Medical Center (STUDY00145615). All caregiver participants gave written informed consent, as did those PWD who were determined to be able to respond on their own via questions on comprehension of the informed consent. This study was registered in ClinicalTrials.gov under the ID NCT04418232.

### Intervention

Alianza Latina is a six-month bilingual intervention, designed to cater to the unique needs of Latino caregivers. It integrates *CuidaTEXT*, an SMS program, with monthly phone consultations facilitated by a trained coach from the research team. These consultations aim to identify unaddressed needs and provide necessary support by brainstorming about different situations that may happen to the caregiver, reinforcing content and skills covered by the text message libraries, and assisting with navigation of community resources. Participants in *CuidaTEXT* receive scheduled messages and can also request on-demand assistance by texting. The intervention, and its development process, have been extensively detailed previously (Perales-Puchalt et al., 2021). *CuidaTEXT* entails sending 1-3 automated daily messages covering various aspects such as logistics, dementia education, self-care, social support, end-of-life care, managing dementia-related behaviors, problem-solving strategies, and community resources. Additionally, participants can text keyword-based queries for immediate assistance on the above-mentioned topics and engage in live chat sessions with the coach for further guidance upon request.

### Assessment

The research team collected information from three sources: baseline survey, six-month follow-up survey, and metrics of text message and phone call interactions. We collected sociodemographic, acculturation and care-related information from the caregivers and PWD at baseline.

Outcomes included feasibility, acceptability, and preliminary efficacy:

#### Feasibility outcomes

were similar to those reported in our previous study (Perales-Puchalt et al.). Outcomes included the feasibility of enrollment (enrollment duration and participants screened), of intervention enrollment (percentage of participants who were able to opt into Alianza Latina), retention rate (completing follow-up survey), assessment (completing preliminary efficacy outcomes), intervention delivery (technical issues), and intervention engagement (usage of texting and phone call features, unsubscribing, and reports of compliance with Alianza Latina).

#### Acceptability outcomes

were also like the ones used in our previous study (Perales-Puchalt et al.) We collected all of them in the six-month follow-up survey. These include six questions: The first four questions asked to what extent Alianza Latina had provided support, helped manage dementia-related changes, helped understand dementia more, and helped in the disease process. These four questions had four response options ranging from not at all to a lot. A fifth question asked about the overall satisfaction with Alianza Latina with four response options ranging from not at all to extremely. The last question asked whether participants would recommend Alianza Latina (yes vs no).

#### Preliminary efficacy outcomes

included four scales about the caregiver and three about the PWD. Caregiver scales included the Neuropsychiatric Inventory Questionnaire (NPI-Q) distress scale (score 0 to 60 distress units) (Kaufer et al., 2000), Six Item Zarit Burden Interview (ZBI-6; scores 0 to 24 burden units) (Higginson et al., 2010), 10 Item Center for Epidemiological Studies Depression scale (CES-D-10; scores: 0 to 30 depressive symptom units) (Andresen et al., 1994), and the One Item Satisfaction with Life Scale (scores: 1-very dissatisfied to 4-very satisfied) (Cheung & Lucas, 2014). PWD scales included the NPI-Q severity scale (score 0 to 36 severity units), Geriatric Depression Scale (GDS; score 0 to 30 depressive symptom units) (Sheikh et al., 1986), and Quality of Life in Alzheimer’s Disease (QOL-AD; score 13 to 52 quality of life units) (Logsdon et al., 2002). Caregivers completed all scales except the GDS and QOL-AD, which were completed by the PWD in the 12 instances where the PWD was able to consent. We reported these scales’ characteristics and their good psychometric properties in the current US Latino sample previously (Perales-Puchalt et al.). The research team administer all these scales at baseline and follow-up.

### Analysis

We used descriptive statistics to summarize the baseline characteristics of both caregivers and persons with dementia (PWD), as well as to report feasibility and acceptability outcomes. To assess preliminary efficacy, we conducted paired-samples t-tests to evaluate changes from baseline to follow-up, as all score distributions were normally distributed. We report means at baseline and follow-up assessments, mean differences between these two points and their 95% confidence intervals. For these t-tests, we also report effect sizes using Cohen’s d’s thresholds of small (0.2), medium (0.5), and large (0.8) (Cohen, 2013). We performed all analyses using SPSS Version 22 (IBM Corp., 2023). The significance level was set at p < 0.05.

## Results

Table 1 summarizes the demographic and background characteristics of Alianza Latina participants. Twenty-six caregivers were part of 10 clusters of 2 (n = 5), 3 (n = 4), and 4 (n = 1) caregivers of the same PWD. Caregivers had a mean age of 52.0 years (SD = 10.2). The majority identified as Latino (95.0%), with 82 identifying as women (82.0%). Forty-three caregivers (43.0%) were born in Mexico, while 30 (30.0%) were born in other Latin American countries. Spanish was the primary language for 55 caregivers (55.0%). Most were either married or living with a partner (72.0%), and nearly one-third (31.0%) reported difficulty or great difficulty covering basic expenses. A large portion (73.0%) were adult children of the PWD.

**Table 1.** Baseline characteristics of the participants enrolled in *Alianza Latina*.

| Caregivers | Total<br>(n=100) | Completers<br>(n=88) | Completers<br>(n=12) |
| --- | --- | --- | --- |
| Age in years, mean (SD) | 52.0 (10.2) | 52.4 (9.6) | 49.3 (13.6) |
| Women, % (n) | 82.0% (82) | 81.8% (72) | 83.3% (10) |
| Latino ethnicity, % (n) | 95.0% (95) | 96.6% (84) | 91.7% (11) |
| Race, % (n) |  |  |  |
| Other, % (n) | 71.0% (71) | 69.3% (61) | 83.3% (10) |
| White, % (n) | 25.0% (25) | 26.1% (23) | 16.7% (2) |
| Multiple races, % (n) | 2.0% (2) | 2.3% (2) | 0.0% (0) |
| Black, % (n) | 1.0% (1) | 1.1% (1) | 0.0% (0) |
| Don't know, % (n) | 1.0% (1) | 1.1% (1) | 0.0% (0) |
| Country of birth |  |  |  |
| US, % (n) | 27.0% (27) | 26.1% (23) | 33.3% (4) |
| Mexico, % (n) | 43.0% (43) | 44.3% (39) | 33.3% (4) |
| Other, % (n) | 30.0% (30) | 29.5% (26) | 33.3% (4) |
| Years in the US among those born abroad, m (SD) | 28.8 (14.2) | 29.1 (14.1) | 27.0 (16.0) |
| USA region or residence |  |  |  |
| Midwest, % (n) | 70.0% (70) | 71.6% (63) | 58.3% (7) |
| West, % (n) | 21.0% (21) | 18.2% (16) | 41.7% (5) |
| South, % (n) | 6.0% (6) | 6.8% (6) | 0.0% (0) |
| East, % (n) | 3.0% (3) | 3.4% (3) | 0.0% (0) |
| Years of education, m (SD) | 12.6 (4.4) | 12.4 (4.4) | 13.5 (4.1) |
| Without medical insurance, % (n) | 34.0% (34) | 33.0% (29) | 41.7% (5) |
| No regular source of medical care, % (n) | 17.0% (17) | 15.9% (14) | 25.0% (3) |
| Spanish only as primary language, % (n) | 55.0% (55) | 55.7% (49) | 50.0% (6) |
| Married or have a partner, % (n) | 72.0% (72) | 71.6% (63) | 81.8% (9) |
| Currently working, % (n) | 64.0% (64) | 67.0% (59) | 41.7% (5) |
| Difficult or very difficult to pay for basic needs, % (n) | 31.0% (31) | 30.7% (27) | 33.3% (4) |
| Relation to PWD |  |  |  |
| Children, % (n) | 73.0% (73) | 71.6% (63) | 66.7% (8) |
| Partner, % (n) | 11.0% (11) | 10.2% (9) | 16.7% (2) |
| Children-in-law, % (n) | 4.0% (4) | 4.5% (4) | 0.0% (0) |
| Other, % (n) | 12.0% (12) | 11.3% (10) | 16.7% (2) |
| PWD | Total (n=84) | Completers<br>(n=73) | Non-completers<br>(n=11) |
| Age in years, m (SD) | 78.2 (10.2) | 78.5 (10.0) | 75.8 (12.0) |
| Woman, % (n) | 70.2% (59) | 71.2% (52) | 63.6% (7) |
| Latino ethnicity, % (n) | 100.0% (84) | 100.0% (73) | 100.0% (11) |
| Race, % (n) |  |  |  |
| Other, % (n) | 81.0% (68) | 80.8% (59) | 81.8% (9) |
| White, % (n) | 17.9% (15) | 17.8% (13) | 18.2% (2) |
| Black, % (n) | 1.2% (1) | 1.4% (1) | 0.0% (0) |
| Country of birth |  |  |  |
| US, % (n) | 11.9% (10) | 12.3% (9) | 9.1% (1) |
| Mexico, % (n) | 53.6% (45) | 53.4% (39) | 54.5% (6) |
| Other, % (n) | 34.5% (29) | 34.2% (25) | 36.4% (4) |
| Years in the US among those born abroad, m (SD) | 36.9 (20.9) | 38.5 (21.3) | 27.0 (16.1) |
| Years of education, m (SD) | 11.2 (20.4) | 11.8 (21.7) | 7.4 (5.1) |
| Without medical insurance, % (n) | 8.3% (7) | 8.2% (6) | 9.1% (1) |
| No regular source of medical care, % (n) | 7.1% (6) | 8.2% (6) | 0.0% (0) |
| Spanish only as primary language, % (n) | 84.5% (71) | 84.9% (62) | 81.8% (9) |
| Married or have a partner, % (n) | 32.2% (27) | 30.1% (22) | 45.5% (5) |
| Currently working, % (n) | 1.2% (1) | 1.4% (1) | 0.0% (0) |

PWD had an average age of 78.2 years (SD = 10.2) and all identified as Latino. Among them, 59 (70.2%) were women, 45 (53.6%) were born in Mexico, and 29 (34.5%) were born in other Latin American countries. Spanish was the main language for 71 PWD (84.5%), and 27 (32.2%) were married or cohabiting. Among the 84 PWD, diagnoses included Alzheimer’s disease (45.2%), unspecified dementia (32.1%), vascular dementia (6.0%), Lewy Body or Parkinson’s-related dementia (6.0%), mixed dementia (4.8%), frontotemporal dementia (2.4%), other types (2.4%), and mild cognitive impairment (1.2%).

Caregivers who completed the study were more often born in Mexico, lived in the Midwest, had access to health insurance and a usual source of care, were not married or partnered, were employed, and tended to be adult children of the PWD. PWD who completed the study had more years of formal education and were less likely to be married or living with a partner.

Table 2 shows the feasibility and acceptability outcomes.

**Table 2.**
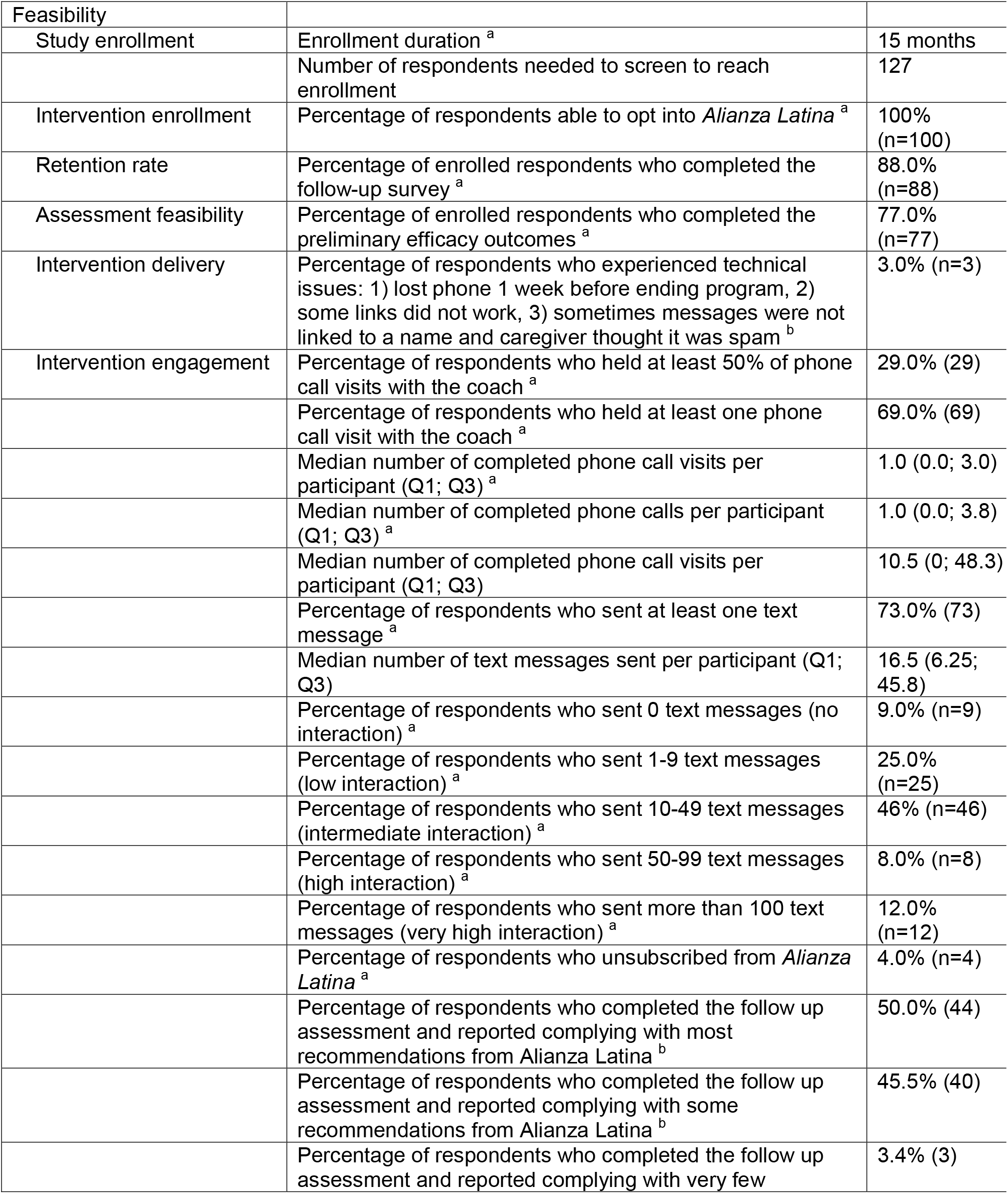
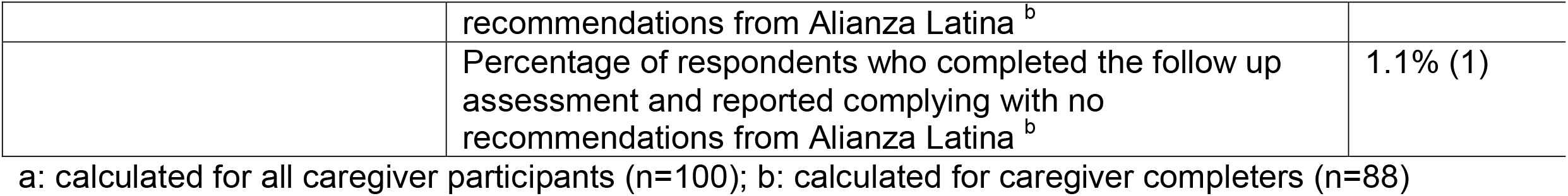
Feasibility of *Alianza Latina* a: calculated for all caregiver participants (n=100); b: calculated for caregiver completers (n=88)

The duration of enrollment was 15 months, and the number of respondents needed to screen to reach full enrollment was 127. The reasons for screen failure included lack of diagnosis (n=3), not attending a primary care clinic (n=1), and no longer being able or willing to participate (n=23). Among the 100 enrolled caregivers, all enrolled (received the initial *CuidaTEXT* message) without any technical issues such as *CuidaTEXT* messages being blocked by the phone carrier, 88 (88.0%) completed at least the acceptability questions of the follow-up survey, and 77 (77.0%) completed the whole survey including the efficacy outcomes. The reason why 11 participants completed the acceptability questions but not the efficacy outcomes was that six PWD died before the follow up assessment (unrelated to their participation). Despite this, no unanticipated problems took place during the study. We did not collect the efficacy information for these participants because it would not reflect the effect of the intervention. Among the 100 enrolled caregivers, three (3.0%) reported experiencing technical issues during their participation, which included losing their phone one week before ending the program, some of the text links not working, and sometimes their first name did not appear in the text messages and the caregiver thought it was spam. Sixty-nine caregivers (69.0%) held at least one monthly phone call visit with the coach, and 29% completed at least 50% of the six-monthly phone call visits. Sixty-six (66.0%) participants sent 10 or more text messages (intermediate-very high interaction), and four (4.0%) unsubscribed from the intervention. Most participants (95.5%) reported complying with most or some recommendations provided by Alianza Latina.

Figure 1 shows the distribution of acceptability outcomes among the 88 caregivers who completed the follow-up assessment. The percentage of participants who reported “somehow” or ‘a lot” levels of acceptability of Alianza Latina was 91.0% for providing support, 89.8% for helping manage disease challenges, 92.0% for helping understand dementia more, 87.9% for helping in the disease process. The percentage of participants who reported “very” or ‘extremely” levels of satisfaction with the intervention was 95.0%, and all respondents who completed the follow up assessment (n=88) said they would recommend Alianza Latina to a friend.

**Figure 1.**
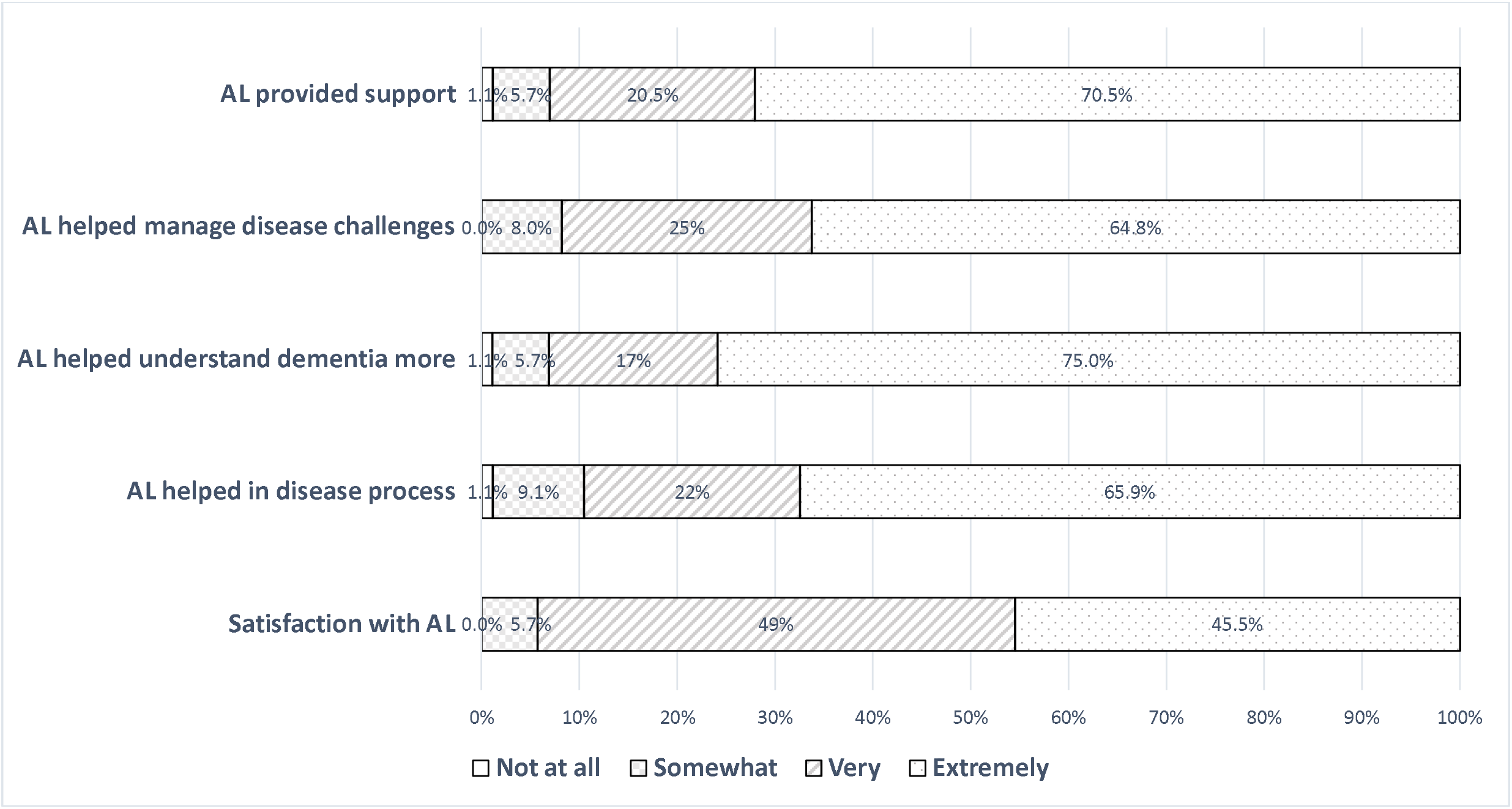
Acceptability of Alianza Latina (n=88)

Table 3 shows the preliminary efficacy of *Alianza Latina* on caregiver and PWD outcomes. When considering the full sample, no change in outcomes was statistically significant, and only depressive symptoms among PWD achieved a clinically significant effect (Cohen’s d=0.2; small). Effect sizes of change in all preliminary efficacy outcomes increased gradually as we restricted samples to those with worse baseline outcome values. In fact, all changes achieved a large effect size if restricted to sample sizes with very poor levels of those outcomes (Cohen’s d=0.73 to 1.59) except PWD’s depressive symptoms (Cohen’s d=0.63; medium) and quality of life (Cohen’s d=-0.22; small). Changes reached statistical significance with small to medium effect sizes in four outcomes when restricted to sample sizes with slightly poorer levels than the total sample (caregiver distress, depressive symptoms and satisfaction with life, and PWD’s depressive symptoms).

**Table 3.**
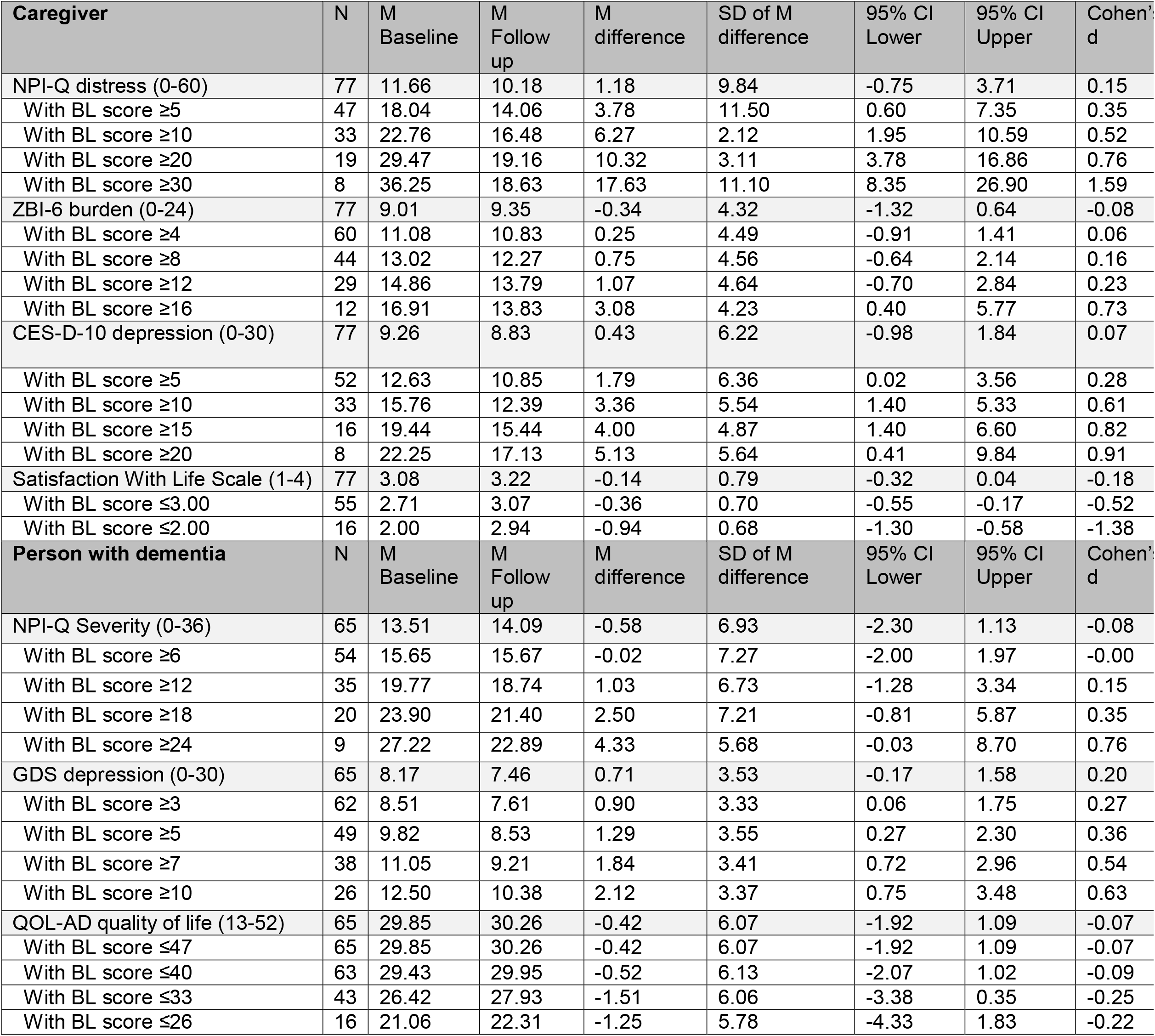
Preliminary efficacy outcomes comparing pre- and post-intervention scores.

| Caregiver | N | M Baseline | M Follow up | M difference | SD of M difference | 95% CI Lower | 95% CI Upper | Cohen's d |
| --- | --- | --- | --- | --- | --- | --- | --- | --- |
| NPI-Q distress (0-60) | 77 | 11.66 | 10.18 | 1.18 | 9.84 | -0.75 | 3.71 | 0.15 |
| With BL score $\geq 5$ | 47 | 18.04 | 14.06 | 3.78 | 11.50 | 0.60 | 7.35 | 0.35 |
| With BL score $\geq 10$ | 33 | 22.76 | 16.48 | 6.27 | 2.12 | 1.95 | 10.59 | 0.52 |
| With BL score $\geq 20$ | 19 | 29.47 | 19.16 | 10.32 | 3.11 | 3.78 | 16.86 | 0.76 |
| With BL score $\geq 30$ | 8 | 36.25 | 18.63 | 17.63 | 11.10 | 8.35 | 26.90 | 1.59 |
| ZBI-6 burden (0-24) | 77 | 9.01 | 9.35 | -0.34 | 4.32 | -1.32 | 0.64 | -0.08 |
| With BL score $\geq 4$ | 60 | 11.08 | 10.83 | 0.25 | 4.49 | -0.91 | 1.41 | 0.06 |
| With BL score $\geq 8$ | 44 | 13.02 | 12.27 | 0.75 | 4.56 | -0.64 | 2.14 | 0.16 |
| With BL score $\geq 12$ | 29 | 14.86 | 13.79 | 1.07 | 4.64 | -0.70 | 2.84 | 0.23 |
| With BL score $\geq 16$ | 12 | 16.91 | 13.83 | 3.08 | 4.23 | 0.40 | 5.77 | 0.73 |
| CES-D-10 depression (0-30) | 77 | 9.26 | 8.83 | 0.43 | 6.22 | -0.98 | 1.84 | 0.07 |
| With BL score $\geq 5$ | 52 | 12.63 | 10.85 | 1.79 | 6.36 | 0.02 | 3.56 | 0.28 |
| With BL score $\geq 10$ | 33 | 15.76 | 12.39 | 3.36 | 5.54 | 1.40 | 5.33 | 0.61 |
| With BL score $\geq 15$ | 16 | 19.44 | 15.44 | 4.00 | 4.87 | 1.40 | 6.60 | 0.82 |
| With BL score $\geq 20$ | 8 | 22.25 | 17.13 | 5.13 | 5.64 | 0.41 | 9.84 | 0.91 |
| Satisfaction With Life Scale (1-4) | 77 | 3.08 | 3.22 | -0.14 | 0.79 | -0.32 | 0.04 | -0.18 |
| With BL score $\leq 3.00$ | 55 | 2.71 | 3.07 | -0.36 | 0.70 | -0.55 | -0.17 | -0.52 |
| With BL score $\leq 2.00$ | 16 | 2.00 | 2.94 | -0.94 | 0.68 | -1.30 | -0.58 | -1.38 |
| Person with dementia | N | M Baseline | M Follow up | M difference | SD of M difference | 95% CI Lower | 95% CI Upper | Cohen's d |
| NPI-Q Severity (0-36) | 65 | 13.51 | 14.09 | -0.58 | 6.93 | -2.30 | 1.13 | -0.08 |
| With BL score $\geq 6$ | 54 | 15.65 | 15.67 | -0.02 | 7.27 | -2.00 | 1.97 | -0.00 |
| With BL score $\geq 12$ | 35 | 19.77 | 18.74 | 1.03 | 6.73 | -1.28 | 3.34 | 0.15 |
| With BL score $\geq 18$ | 20 | 23.90 | 21.40 | 2.50 | 7.21 | -0.81 | 5.87 | 0.35 |
| With BL score $\geq 24$ | 9 | 27.22 | 22.89 | 4.33 | 5.68 | -0.03 | 8.70 | 0.76 |
| GDS depression (0-30) | 65 | 8.17 | 7.46 | 0.71 | 3.53 | -0.17 | 1.58 | 0.20 |
| With BL score $\geq 3$ | 62 | 8.51 | 7.61 | 0.90 | 3.33 | 0.06 | 1.75 | 0.27 |
| With BL score $\geq 5$ | 49 | 9.82 | 8.53 | 1.29 | 3.55 | 0.27 | 2.30 | 0.36 |
| With BL score $\geq 7$ | 38 | 11.05 | 9.21 | 1.84 | 3.41 | 0.72 | 2.96 | 0.54 |
| With BL score $\geq 10$ | 26 | 12.50 | 10.38 | 2.12 | 3.37 | 0.75 | 3.48 | 0.63 |
| QOL-AD quality of life (13-52) | 65 | 29.85 | 30.26 | -0.42 | 6.07 | -1.92 | 1.09 | -0.07 |
| With BL score $\leq 47$ | 65 | 29.85 | 30.26 | -0.42 | 6.07 | -1.92 | 1.09 | -0.07 |
| With BL score $\leq 40$ | 63 | 29.43 | 29.95 | -0.52 | 6.13 | -2.07 | 1.02 | -0.09 |
| With BL score $\leq 33$ | 43 | 26.42 | 27.93 | -1.51 | 6.06 | -3.38 | 0.35 | -0.25 |
| With BL score $\leq 26$ | 16 | 21.06 | 22.31 | -1.25 | 5.78 | -4.33 | 1.83 | -0.22 |

## Discussion

In this study, we aimed to test the feasibility, acceptability and preliminary efficacy of Alianza Latina, a combination of an SMS texting intervention (CuidaTEXT) plus optional monthly phone calls among mostly Latino informal caregivers and the Latino PWD they care for. In line with CuidaTEXT findings, most aspects of the Alianza Latina study design and intervention are highly feasible and caregiver participants are highly satisfied with the intervention. The intervention led to improved outcomes among caregivers and PWD only when restricting the sample to those who had at least a minimal level of baseline severity in the outcome studied.

Our findings show that the Alianza Latina study design is feasible. Results align with our CuidaTEXT study, were we tested the feasibility of an SMS texting intervention among Latino caregivers (Perales-Puchalt, Ramírez-Mantilla, et al., 2022). For example, the number of people needed to screen to achieve the sample was 27%, and retention rate was 88% in this study vs 29% and 87% in the CuidaTEXT study. Other similarities include the lack of issues with opting into the intervention, very low number of technical issues, and the very low percentage of participants who unsubscribed from the intervention. The low percentage of Alianza Latina participants unsubscribing is promising even when comparing to feasible and preliminary efficacious SMS texting interventions for other conditions such as smoking cessation. For example, in the QuitNowTXT study in Israel, 34% of smokers unsubscribed by the 4^th^ week (Abroms et al., 2015). Levels of acceptability with the intervention were also similar to those of CuidaTEXT and higher than other texting interventions such as QuitNowTXT (Abroms et al., 2015). The levels of feasibility and acceptability are likely a product of careful co-design with the target population (Perales-Puchalt, Acosta-Rullán, et al., 2022).

A notable difference with the CuidaTEXT feasibility study is that engagement with texting was lower (e.g., 12% sent more than 100 texts vs 25%), perhaps due to the additional phone call modality option. Our previous findings show that the frequency of texting in CuidaTEXT is not associated with clinical outcomes (Medina et al., 2023). Participants completed an average of one out of six possible phone call visits. The lower use of phone calls compared to texting is in line with the low acceptability of phone call-based interventions among Latinos (Chodosh et al., 2015). We plan to analyze the association between phone call engagement and feasibility and clinical outcomes in a future publication.

Our clinical outcomes in the overall sample did not achieve the promising effects as the CuidaTEXT feasibility study (Perales-Puchalt, Ramírez-Mantilla, et al., 2022). While effect sizes were generally smaller than the CuidaTEXT study, baseline levels of most clinical outcomes were better in the current study (e.g., 12 vs 20 out of 60 for caregiver distress). Improvement in preliminary efficacy outcomes were only evident when restricting the sample to those with at least a minimum level of severity. This increase in clinical significance applied to three caregiver outcomes (distress, depressive symptoms and satisfaction with life) and one outcome of the PWD (depressive symptoms), which reached effect sizes ranging from small to large, depending on the outcome and the amount of sample restriction. In fact, restricting the sample to those with NPI-Q distress scores ≥5 led to a mean difference of 3.8, which is above the 3.1 reported to be minimally clinically significant with this scale (Mao et al., 2015). These findings suggest that a future Alianza Latina trial needs to include its primary outcomes in the eligibility criteria and apply the cutoff that most apply to the effect size it intends to achieve compared to their control group. For example, if a future trial aims to detect a medium PWD depressive symptom effect of Alianza Latina compared to a control group that is not expected to change this outcome, eligibility should restrict the sample to only those with GDS scores ≥5.

This study has limitations. First, this was a feasibility trial and did not include a control group or randomization. Therefore, changes in clinical outcomes are preliminary, and not necessarily caused by the intervention. Our inability to follow up with 12 participants has prevented us from understanding their reasons not to complete follow-up assessments, their opinions about Alianza Latina or its preliminary efficacy. Despite the relatively large sample size for a feasibility study and its diversity (e.g., different primary language, US regions, countries of birth), the sample was non-probabilistic and male caregivers were under-represented, which reduces the external validity of these findings. Intervention masking was not possible due to its behavioral nature, which could bias responses. We did not analyze the participants’ quantitative and qualitative interactions with the text message intervention in depth, as this is beyond the scope of the current manuscript. Future studies will analyze these interactions and their association with outcomes, to shed further light on mechanisms of action.

This intervention, which combines SMS texting with optional monthly phone support, holds important implications for public health, clinical care, and future research. Its potential for broad implementation is bolstered by the widespread availability of text messaging and its lower-than usual reliance on workforce resources—features that contribute to greater cost-effectiveness and scalability. Findings from this study support the importance of designing interventions with implementation in mind from the outset, emphasizing user-centered approaches that align with real-world needs and settings (Gaugler et al., 2021; International Organization for Standardization, 2018).

Moving forward, the most appropriate next step is to test the intervention’s efficacy through a randomized controlled trial aligned with Stage 2 of the NIH Stage Model for Behavioral Intervention Development (Onken et al., 2014). In fact, we are currently conducting a randomized controlled trial testing the efficacy of the texting component (CuidaTEXT) among 288 Latino family caregivers. Informed by this and prior research (Perales-Puchalt, Ramírez-Mantilla, et al., 2022), we are limiting the sample to those with CES-D-10 scores≥ 7. If found effective, the intervention could be integrated into clinical and community settings with ease, allowing caregivers to enroll via text or enabling staff to register them through a simple web platform. Future trials should also aim to examine whether there is value in adding optional phone calls and potential mechanisms of action suggested by the current study.

## Conclusion

The current study shows that combining SMS texting with phone calls for dementia caregiver support is feasible and acceptable. Testing the efficacy of this intervention will require restricting the sample to those with at least minimal severity in caregiver distress, depressive symptoms and quality of life, or PWD depressive symptoms. This study adds to the suboptimal literature on ways to increase access and improve mental health outcomes among Latino caregivers of people with dementia and PWD, who are understudied (Dessy et al., 2022), and disproportionately impacted (Chen et al., 2020; Salazar et al., 2017).

## Data Availability

Data produced in the present study are available upon reasonable request to the authors

## Acknowledgements

JPP thanks the national and local organizations that have partnered with him to conduct present and past research since 2015. The research team thanks research participants included in all stages of this research as well as anyone who has contributed directly and indirectly to this research. We also thank Visión y Compromiso, the UsAgainstAlzheimer’s A-List and the Alzheimer’s Association’s TrialMatch for sharing the opportunity to participate with the people they serve. The ideas and opinions expressed herein are those of the authors alone, and endorsement by the authors’ institutions or the funding agency is not intended and should not be inferred.

